# Out-of-Pocket Spending for COVID-19 Hospitalizations in 2020

**DOI:** 10.1101/2021.05.26.21257879

**Authors:** Kao-Ping Chua, Rena M. Conti, Nora V. Becker

**Author notes:** **Address correspondence to:** Kao-Ping Chua, MD, PhD, 300 North Ingalls St, SPC 5456, Room 6E18, Ann Arbor, Michigan 48109-5456.

## Abstract

**IMPORTANCE:** Many insurers waived cost-sharing for COVID-19 hospitalizations during 2020. Nonetheless, patients may have been billed if their plans did not implement waivers or if waivers did not capture all hospitalization-related care, including clinician services. Assessing out-of-pocket spending for COVID-19 hospitalizations in 2020 could demonstrate the financial burden patients may face if insurers allow waivers to expire, as many chose to do during early 2021.

**OBJECTIVE:** To estimate out-of-pocket spending for COVID-19 hospitalizations in 2020

**DESIGN:** Cross-sectional analysis

**SETTING:** IQVIA PharMetrics® Plus for Academics Database, a national claims database

**PARTICIPANTS:** COVID-19 hospitalizations for privately insured and Medicare Advantage patients during March-September 2020

**MAIN OUTCOMES/MEASURES:** Mean total out-of-pocket spending, defined as the sum of out-of-pocket spending for facility services billed by hospitals (e.g., accommodation charges) and for professional/ancillary services billed by clinicians and ancillary providers (e.g., clinician inpatient evaluation and management, ambulance transport)

**RESULTS:** Analyses included 4,075 hospitalizations. Of the 1,377 hospitalizations for privately insured patients and the 2,698 hospitalizations for Medicare Advantage patients, 981 (71.2%) and 1,324 (49.1%) had out-of-pocket spending for facility services, professional/ancillary services, or both. Among these hospitalizations, mean (SD) total out-of-pocket spending was $788 (1,411) and $277 (363). In contrast, 63 (4.6%) and 36 (1.3%) hospitalizations had out-of-pocket spending for facility services. Among these hospitalizations, mean total out-of-pocket spending was $3,840 (3,186) and $1,536 (1,402). Total out-of-pocket spending exceeded $4,000 for 2.5% of privately insured hospitalizations, compared with 0.2% of Medicare Advantage hospitalizations.

**CONCLUSIONS:** Few COVID-19 hospitalizations in this study had out-of-pocket spending for facility services, suggesting most were covered by insurers with cost-sharing waivers. However, many hospitalizations had out-of-pocket spending for professional/ancillary services. Overall, 7 in 10 privately insured hospitalizations and half of Medicare Advantage hospitalizations had any out-of-pocket spending. Findings suggest insurer cost-sharing waivers may not cover all hospitalization-related care. Moreover, high cost-sharing for some hospitalizations suggests out-of-pocket burden could be substantial if waivers expire, particularly for privately insured patients. Rather than rely on voluntary insurer actions to mitigate this burden, federal policymakers should consider mandating insurers to waive cost-sharing for all COVID-19 hospitalization-related care throughout the pandemic.

**KEY POINTS:** *Question:* How much were patients billed for COVID-19 hospitalizations in 2020?

*Findings:* Of 1,377 and 2,698 COVID-19 hospitalizations for privately insured and Medicare Advantage patients, 71.2% and 49.1% had cost-sharing for facility services billed by hospitals, services billed by clinicians or ancillary providers, or both. Among these hospitalizations, mean out-of-pocket spending was $788 and $277. 4.6% and 1.3% had cost-sharing for facility services; among these hospitalizations, mean out-of-pocket spending was $3,840 and $1,536.

*Meaning:* Insurer cost-sharing waivers for COVID-19 hospitalizations may not cover all hospitalization-related care. Patient out-of-pocket burden could be substantial if insurers allow waivers to expire.

## INTRODUCTION

Between August 2020 and April 2021, there were more than 2.1 million U.S. hospitalizations for coronavirus disease 2019 (COVID-19).^1^ To mitigate patient financial burden, many private insurers and Medicaid Advantage insurers voluntarily waived cost-sharing for COVID-19 hospitalizations during part or all of 2020.^2,3^ Literature examining cost-sharing for other respiratory infection hospitalizations suggests these waivers potentially resulted in substantial savings for patients.^4-6^ For example, among privately insured patients hospitalized for respiratory infections between 2016-2019, average out-of-pocket spending was $1,653 for patients in traditional plans and $1,961 for patients in consumer-driven health plans.^4^ Among Medicare Advantage patients hospitalized for influenza in 2018, average out-of-pocket spending was almost $1,000.^6^

While waivers may have mitigated financial burden for many patients hospitalized for COVID-19 during 2020, some patients may still have been billed if their plans did not implement waivers or if waivers did not capture all hospitalization-related care. Hospitalizations can result in two categories of bills.^7,8^ The first includes facility services provided by hospitals, such as accommodation and inpatient pharmacy services. The second includes services from clinicians and ancillary providers (hereafter referred to as “professional/ancillary services”). This category includes clinician services for emergency department and inpatient care, as well as ambulance services for transport to the hospital. While waivers would ideally cover both categories, some may only have covered facility services billed by hospitals, not professional/ancillary services billed separately by providers.

Protecting patients from the costs of COVID-19 hospitalizations is an important policy goal, as hospitalization surges may still occur despite vaccination efforts. Despite this, no study to our knowledge has assessed the amount patients were billed for COVID-19 hospitalizations during 2020, either overall or by service category. Addressing this knowledge gap may inform policy in several ways. First, it could motivate efforts to improve the comprehensiveness and implementation of existing insurer cost-sharing waivers for COVID-19 hospitalizations. Second, it could demonstrate the potential financial burden patients may face if insurers allow cost-sharing waivers to expire, as many chose to do during early 2021.^9,10^ Finally, it could illustrate the potential need for federal legislation mandating U.S. insurers to waive cost-sharing for these hospitalizations – legislation that was proposed, but not passed, in the U.S. House of Representatives in 2020.^11^ In this study, we used national claims data to estimate out-of-pocket spending for COVID-19 hospitalizations during March-September 2020 among patients covered by private and Medicare Advantage plans.

## METHODS

### Data source

In May 2021, we conducted a cross-sectional analysis of the IQVIA PharMetrics^®^ Plus for Academics database. This database contains fully-adjudicated medical and pharmacy claims from de-identified patients in all 50 states and the District of Columbia. Claims were complete through September 30, 2020 when data were delivered at the end of March 2021, corresponding to a six-month claims run-off period. The database included 1.0 million patients covered by Medicare Advantage plans and 7.7 million patients covered by fully-insured private plans in 2020. The database did not include any patients covered by self-insured private plans.

Data include patient year of birth, state and region of residence, payer type, and plan type. Data also include International Classification of Diseases, Tenth Revision, Clinical Modification (ICD-10-CM) diagnosis codes, a hospitalization identifier assigned to all claims that occurred on or between the admission and discharge dates of hospitalizations, amounts billed to patients (deductibles, co-insurance, and co-payments), and a flag for whether the billing provider was a hospital, clinician, or other entity. The database does not include information on race, ethnicity, household income, out-of-network status, or in-hospital death (to protect patient confidentiality). Moreover, the database does not include plan identifiers or information on plan benefit design, including whether insurers had cost-sharing waivers for COVID-19 hospitalizations. As discussed below, we conducted analyses to evaluate whether such waivers may have been in place. Because data were de-identified, the Institutional Review Board of the University of Michigan Medical School exempted analyses from human subjects review.

### Study sample

We included hospitalizations that had a primary diagnosis of confirmed COVID-19 infection (ICD-10-CM diagnosis code U071) and that began and ended between March 1-September 29, 2020. We required discharge before September 30, 2020 to ensure the end of hospitalizations was observed (see **Appendix 1** for details). We excluded hospitalizations if they were covered by a secondary insurer (e.g., a different private insurance plan) or if any associated claim had missing data for out-of-pocket spending or billing provider type.

### Categorization of claims

For each hospitalization, we assigned claims with the corresponding hospitalization identifier to 1 of 3 mutually exclusive categories (see **Appendix 2** for details):

1. Claims for facility services (institutional claims with a hospital or emergency department place of service and a hospital billing provider type). These services included but were not limited to hospital accommodation, facility charges for emergency department visits, and inpatient laboratory, pharmacy, and radiology services.
2. Claims for professional/ancillary services, defined as one of three types of services:
  - Ambulance (claims with an ambulance place of service or procedure code)
  - Clinician (claims with an emergency department or hospital place of service and clinician billing provider type)
  - Miscellaneous (claims with billing provider type for miscellaneous providers, such as durable medical equipment providers and dialysis centers).
  For additional context, clinician services were divided into 4 subtypes:
  - Emergency department (claims with an emergency department place of service)
  - Inpatient evaluation and management (claims with a hospital place of service and procedure code for evaluation and management, e.g., initial or subsequent hospital care)
  - Inpatient diagnostic testing (claims with hospital place of service and procedure codes for laboratory tests, radiology tests, electrocardiograms, echocardiography, electroencephalograms, and vascular diagnostic studies)
  - Other inpatient services (claims with hospital place of service and procedure codes for services other than evaluation and management and diagnostic testing, such as procedures).
3. Unclassified claims. This category included the approximately 4.3% of claims that were assigned the confinement identifier for the COVID-19 hospitalization but did not meet criteria for a facility or professional/ancillary service. Three-quarters of these claims had a place of service code for office, home, or hospital outpatient department. While some of these claims could represent care at visits resulting in direct admission to the hospital, they could also include care provided at unrelated visits. In the main analysis, we excluded these claims to maximize the probability of only capturing out-of-pocket spending for services truly associated with hospitalizations. We included these claims in a sensitivity analysis.

### Outcomes

Out-of-pocket spending was defined as the sum of deductibles, co-insurance, and co-payments. For each payer type (private insurance and Medicare Advantage), we determined the proportion of hospitalizations in two categories: those with out-of-pocket spending for facility services (with or without out-of-pocket spending for professional/ancillary services), and those with out-of-pocket spending for facility services, professional/ancillary services, or both. For hospitalizations in both categories, we calculated total out-of-pocket spending, defined as the sum of out-of-pocket spending across facility and professional/ancillary services. Additionally, we calculated the proportion of all hospitalizations with out-of-pocket spending for the 3 main types of professional/ancillary services and for the 4 subtypes of clinician services.

### Presence of cost-sharing waivers

The database did not report whether COVID-19 hospitalizations were covered by plans with cost-sharing waivers. However, as noted below, the vast majority of hospitalizations in our sample did not have cost-sharing for facility services. While this might suggest that most hospitalizations were covered by insurers that waived cost-sharing for facility services – that is, that the absence of cost-sharing for facility services implied the presence of a waiver – a potential alternative explanation is that most patients had already met their plan’s annual out-of-pocket maximum at the time of hospitalizations. To evaluate this possibility, we conducted a sensitivity analysis in which we restricted analyses to hospitalizations by patients continuously enrolled since January 2020, calculated out-of-pocket spending across medical and pharmacy claims in 2020 prior to the hospitalization, and calculated the incidence of out-of-pocket spending for facility services among hospitalizations for patients in the lowest quartile of this prior out-of-pocket spending. These patients likely had not met out-of-pocket maximums at the time of hospitalizations. If few of these patients had cost-sharing for facility services, that would support the notion that cost-sharing waivers, rather than meeting out-of-pocket maximums, drove the low observed incidence of cost-sharing for facility services.

We also explored whether it was reasonable to assume that hospitalizations with out-of-pocket spending for facility services were not covered by insurers with cost-sharing waivers for these services (i.e., that the presence of cost-sharing for facility services implied the absence of a waiver – the inverse of the assumption above). To evaluate this assumption, we compared the incidence of out-of-pocket spending for facility services between COVID-19 hospitalizations and influenza hospitalizations. The latter require similar care as COVID-19 hospitalizations, but no insurers to our knowledge waived cost-sharing for influenza hospitalizations during the study period. If the presence of out-of-pocket spending for facility services implies the absence of a waiver for these services, a much higher proportion of influenza hospitalizations would have out-of-pocket spending for facility services compared with COVID-19 hospitalizations. In this analysis, influenza hospitalizations were those that met similar inclusion and exclusion criterion but had a primary diagnosis of influenza (ICD-10-CM diagnosis code J09-J11). No influenza hospitalizations included also had a COVID-19 diagnosis code (U017).

### Statistical analysis

We used descriptive statistics to assess patient characteristics, length of stay, and intensive care unit utilization (based on revenue codes corresponding to accommodation charges for intensive care or coronary care units; **Appendix 2**). To contextualize cost-sharing amounts, we calculated mean and median allowed amounts (reimbursement to providers plus patient liability) across facility and professional/ancillary services among privately insured and Medicare Advantage hospitalizations separately. Analyses used SAS 9.4 (SAS Institute).

## RESULTS

### Sample characteristics

Of 4,371 COVID-19 hospitalizations meeting inclusion criteria, 230 were excluded because the insurer was secondary, 63 because data on billing provider type were missing, and 3 because out-of-pocket spending data were missing. Overall, 296 (6.8%) hospitalizations were excluded, leaving 4,075 hospitalizations. These hospitalizations occurred among 3,875 unique patients; 282 patients had 2 hospitalizations, while 9 patients had 3 hospitalizations.

**Table 1** displays characteristics of the 4,075 hospitalizations. Overall, 1,377 (33.8%) and 2,698 (66.2%) hospitalizations were for privately insured and Medicare Advantage patients. Of the former, 552 (40.1%) were for female patients. Mean length of stay was 7.3 (SD 7.6) days; 640 (46.5%) hospitalizations involved intensive care unit utilization. Of 2,698 hospitalizations for Medicare Advantage patients, 1,432 (53.1%) were for females. Mean length of stay was 9.2 days (SD 8.9); 1,212 (44.9%) hospitalizations involved intensive care unit utilization.

**Table 1.** Characteristics of COVID-19 hospitalizations between March-September 2020, IQVIA PharmMetrics Plus for Academics

|  | Private insurance | Medicare Advantage |
| --- | --- | --- |
| **Number of COVID-19 hospitalizations** | 1,377 | 2,698 |
| **Month of admission** |  |  |
| March | 106 (7.7%) | 236 (8.7%) |
| April | 307 (22.3%) | 917 (34.0%) |
| May | 120 (8.7%) | 331 (12.3%) |
| June | 147 (10.7%) | 200 (7.4%) |
| July | 352 (25.6%) | 413 (15.3%) |
| August | 204 (14.8%) | 392 (14.5%) |
| September | 141 (10.2%) | 209 (7.7%) |
| **Mean length of stay (SD)** | 7.3 (7.6) | 9.2 (8.9) |
| **Any intensive care unit use<sup>a</sup>** | 640 (46.5%) | 1,212 (44.9%) |
| **Age in years** |  |  |
| 0-17 | 12 (0.9%) | 0 (0.0%) |
| 18-25 | 19 (1.4%) | 1 (0.0%) |
| 26-34 | 75 (5.4%) | 7 (0.3%) |
| 35-44 | 201 (14.6%) | 14 (0.5%) |
| 45-54 | 389 (28.2%) | 101 (3.7%) |
| 55-64 | 550 (39.9%) | 285 (10.6%) |
| 65-74 | 107 (7.8%) | 877 (32.5%) |
| 75-85 | 21 (1.5%) | 925 (34.3%) |
| > 85 | 3 (0.2%) | 488 (18.1%) |
| **Sex** |  |  |
| Male | 825 (59.9%) | 1,266 (46.9%) |
| Female | 552 (40.1%) | 1,432 (53.1%) |
| **Region** |  |  |
| Northeast | 200 (14.5%) | 917 (34.0%) |
| Midwest | 315 (22.9%) | 1,062 (39.4%) |
| South | 623 (45.2%) | 433 (16.0%) |
| West | 232 (16.8%) | 274 (10.2%) |
| **Plan type** |  |  |
| Health maintenance organization | 504 (36.6%) | 2,161 (80.1%) |
| Preferred provider organization | 647 (47.0%) | 512 (19.0%) |
| Consumer-directed health plan | 226 (16.4%) | 0 (0.0%) |
| Unknown | 0 (0.0%) | 25 (0.9%) |

<sup>a</sup>Defined as the occurrence of $\geq 1$ claim with a revenue code for intensive care unit or coronary care unit (0200-0209, 0210-0219)

Privately insured hospitalizations were most commonly covered by preferred provider organization plans (47.0%). Mean and median allowed amounts for privately insured hospitalizations was $42,200 (SD 65,328) and $25,339 (25^th^-75^th^ percentile: $16,064-$39,484). Medicare Advantage hospitalizations were most commonly covered by health maintenance organization plans (80.1%). Mean and median allowed amounts for Medicare Advantage hospitalizations were $21,501 (SD 21,387) and $17,480 (25^th^-75^th^ percentile: $14,383-$21,133).

### Out-of-pocket spending

Of the 1,377 and 2,698 hospitalizations for privately insured and Medicare Advantage patients, 63 (4.6%) and 36 (1.3%) had out-of-pocket spending for facility services. Among these 63 and 36 hospitalizations, mean (SD) total out-of-pocket spending was $3,840 (3,186) and $1,536 (1,402). In contrast, of the 1,377 and 2,698 hospitalizations for privately insured and Medicare Advantage patients, 981 (71.2%) and 1,324 (49.1%) had out-of-pocket spending for facility services, professional/ancillary services, or both. Among these 981 and 1,324 hospitalizations, mean total out-of-pocket spending was $788 (1,411) and $277 (363) (**Table 2**). Of all 1,377 privately insured hospitalizations, 99 (7.2%) and 34 (2.5%) had total out-of-pocket exceeding $2,000 and $4,000. Of all 2,698 hospitalizations for Medicare Advantage patients, the corresponding numbers were 7 (0.3%) and 5 (0.2%).

**Table 2.** Incidence and magnitude of out-of-pocket spending among COVID-19 and influenza hospitalizations, IQVIA PharMetrics for Academics Database

| Hospitalization type | No. of hospitalizations (% of total) | Mean (SD) total OOP spending | Mean (SD) OOP spending: facility services<sup>a</sup> | Mean (SD) OOP spending: professional/ancillary services<sup>a</sup> |
| --- | --- | --- | --- | --- |
| **Private insurance, COVID-19 (n = 1,377)** |  |  |  |  |
| Had OOP spending for facility services | 63 (4.6%) | \$3,840 (3,186) | \$3,348 (2,950) | \$492 (849) |
| Had OOP spending for facility services, professional/ancillary services, or both | 981 (71.2%) | \$788 (1,411) | \$215 (1,107) | \$573 (869) |
| **Medicare Advantage, COVID-19 (n = 2,698)** |  |  |  |  |
| Had OOP spending for facility services | 36 (1.3%) | \$1,536 (1,402) | \$1,440 (1,405) | \$97 (147) |
| Had OOP spending for facility services, professional/ancillary services, or both | 1,324 (49.1%) | \$277 (363) | \$39 (327) | \$238 (191) |
| **Private insurance, influenza (n = 61)** |  |  |  |  |
| Had OOP spending for facility services | 51 (83.6%) | \$3,510 (2,524) | \$2,998 (2,293) | \$512 (582) |
| Had OOP spending for facility services, professional/ancillary services, or both | 55 (90.2%) | \$3,327 (2,528) | \$2,780 (2,342) | \$546 (620) |
| **Medicare Advantage, influenza (n = 178)** |  |  |  |  |
| Had OOP spending for facility services | 159 (89.3%) | \$1,226 (708) | \$1,117 (665) | \$109 (144) |
| Had OOP spending for facility services, professional/ancillary services, or both | 173 (97.2%) | \$1,150 (728) | \$1,027 (707) | \$123 (156) |

OOP – out-of-pocket
<sup>a</sup>See Appendix 2 for codes used to identify facility and professional/ancillary services. Facility services were those billed by hospitals for services such as accommodation. Professional/ancillary services were those billed by clinicians and ancillary providers, such as ambulance providers.

**Table 3** shows the incidence and magnitude of out-of-pocket spending for each of the 3 main types of professional/ancillary services and for the 4 subtypes of clinician services. Of the 1,377 hospitalizations for privately insured patients, 137 (9.9%) and 918 (66.7%) had out-of-pocket spending for ambulance services and clinician services. When analyzing clinician services by subtype, 516 (37.5%) and 641 (46.6%) hospitalizations had out-of-pocket spending for inpatient evaluation and management services and diagnostic testing services. Among the 516 hospitalizations with out-of-pocket spending for inpatient evaluation and management services, mean (SD) out-of-pocket spending for these services was $622 (765). Compared with hospitalizations for privately insured patients, hospitalizations for Medicare Advantage patients had a higher incidence of out-of-pocket spending for ambulance services (36.5%) but a lower incidence for clinician services (22.1%).

**Table 3.** Incidence and magnitude of out-of-pocket spending for professional/ancillary services among COVID-19 hospitalizations, IQVIA PharMetrics for Academics Database

| Service type | Privately insured (n = 1,377 hospitalizations) |  |  |  | Medicare Advantage (n = 2,698 hospitalizations) |  |  |  |
| --- | --- | --- | --- | --- | --- | --- | --- | --- |
|  | No. patients with ≥1 claim (% patients in sample) | Mean (SD) OOP spending per patient in sample | No. patients with OOP spending (% patients in sample) | Mean (SD) OOP spending among patients with OOP spending | No. patients with ≥1 claim (% patients in sample) | Mean (SD) OOP spending per patient in sample | No. patients with OOP spending (% patients in sample) | Mean (SD) OOP spending among patients with OOP spending |
| **<u>Main types of professional/ancillary services<sup>a</sup></u>** |  |  |  |  |  |  |  |  |
| Ambulance | 305 (22.1%) | \$59 (248) | 137 (9.9%) | \$596 (550) | 1,425 (52.8%) | \$87 (139) | 985 (36.5%) | \$239 (130) |
| Clinician | 1,334 (96.9%) | \$317 (682) | 918 (66.7%) | \$476 (789) | 2,608 (96.7%) | \$29 (117) | 595 (22.1%) | \$130 (221) |
| Miscellaneous<sup>b</sup> | 272 (19.8%) | \$32 (177) | 167 (12.1%) | \$263 (445) | 401 (14.9%) | \$1 (10) | 99 (3.7%) | \$26 (44) |
| **<u>Subtypes of clinician services</u>** |  |  |  |  |  |  |  |  |
| Emergency department | 746 (54.2%) | \$31 (103) | 399 (29.0%) | \$106 (169) | 1,493 (55.3%) | \$0 (1) | 255 (9.5%) | \$2 (2) |
| Inpatient evaluation and management | 1,234 (89.6%) | \$233 (557) | 516 (37.5%) | \$622 (765) | 2,495 (92.5%) | \$24 (103) | 394 (14.6%) | \$162 (225) |
| Inpatient diagnostic | 668 (48.5%) | \$36 (85) | 641 (46.6%) | \$78 (111) | 1,438 (53.3%) | \$3 (14) | 427 (15.8%) | \$18 (32) |
| Other inpatient<sup>c</sup> | 109 (7.9%) | \$17 (179) | 63 (4.6%) | \$375 (757) | 314 (11.6%) | \$2 (19) | 83 (3.1%) | \$67 (87) |

OOP – out-of-pocket
<sup>a</sup>Professional/ancillary services include those submitted by clinicians and those from ancillary service providers, such as ambulance providers. See Appendix 2 for details
<sup>b</sup>Services from miscellaneous providers, such as a durable medical equipment provider or dialysis center
<sup>c</sup>Includes services submitted by clinicians with a hospital place of service but no procedure code for evaluation and management or diagnostic services (e.g., procedures, anesthesia)

### Analyses assessing presence of cost-sharing waivers

Among hospitalizations for privately insured and Medicare Advantage patients in the lowest quartile of out-of-pocket spending prior to hospitalization, the proportion with out-of-pocket spending for facility services was still modest, at 8.3% and 1.8% **(Appendix 3)**. A total of 61 and 178 influenza hospitalizations for privately insured and Medicare Advantage patients met inclusion and exclusion criterion. Of these hospitalizations, 51 (83.6%) and 159 (89.3%) had out-of-pocket spending for facility services, compared with 4.6% and 1.3% among COVID-19 hospitalizations covered by private insurance and Medicare Advantage plans **(Table 2)**.

### Sensitivity analysis including unclassified claims

When including the 4.3% of claims that did not meet criteria for a facility or professional/ancillary service, the proportion of hospitalizations for privately insured and Medicare Advantage patients with out-of-pocket spending for any associated claim was 73.6% and 53.8%. These proportions were similar to the proportion of hospitalizations with out-of-pocket spending for facility services, professional/ancillary services, or both (71.2% and 49.1%).

## DISCUSSION

In this national study of COVID-19 hospitalizations between March-September 2020, the incidence of out-of-pocket spending differed substantially for facility and professional/ancillary services. When considering facility services only, few COVID-19 hospitalizations had out-of-pocket spending. However, when also considering professional/ancillary services, 7 in 10 privately insured hospitalizations and half of Medicare Advantage hospitalizations had any out-of-pocket spending. If the absence of out-of-pocket spending for facility services is an indicator of the presence of an insurer cost-sharing waiver for these services – an assumption supported by our analyses – then most study hospitalizations were covered by insurers that at least waived cost-sharing for facility services. If true, then the high incidence of out-of-pocket spending for professional/ancillary services suggests that many insurer cost-sharing waivers may fail to capture all hospitalization-related care.

Whether this failure is intentional is unclear. Unlike COVID-19 testing and vaccination, there is no federal mandate for insurers to waive cost-sharing for COVID-19 hospitalizations.^12^ Consequently, insurer waivers could be heterogeneous, with some applying only to facility services and others applying to hospitalization care more broadly. Even if insurers intend for waivers to capture all hospitalization-related care, implementation problems may occur. For example, if insurers do not link clinician inpatient evaluation and management bills to the COVID-19 hospitalization, patients may be billed erroneously.

Insurers and clinicians might consider three steps to mitigate patient financial liability for professional/ancillary services related to COVID-19 hospitalizations. First, insurers with no cost-sharing waiver or with waivers of limited scope could consider implementing a comprehensive waiver, for example one that covers all services on or between the admission and discharge dates of hospitalizations. Second, insurers that already have comprehensive waivers could work to ensure appropriate implementation. Finally, clinicians could encourage patients to contest any bills for professional/ancillary services that should be covered under an insurer’s cost-sharing waiver.

In this study, 4.6% and 1.5% of hospitalizations for privately insured and Medicare Advantage patients had out-of-pocket spending for facility services. Among these hospitalizations, mean total out-of-pocket spending was $3,840 and $1,536, respectively. If the presence of out-of-pocket spending for facility services implies the absence of an insurer cost-sharing waiver for these services – as suggested by the fact that the vast majority of influenza hospitalizations had cost-sharing for facility services – our findings suggest that out-of-pocket burden for COVID-19 hospitalizations could be large without insurer cost-sharing waivers. This would have important policy implications. In early 2021, several large insurers, including Anthem and United Healthcare, allowed their cost-sharing waivers for COVID-19 hospitalizations to expire.^9,10^ Analyses suggest patients covered by these insurers could now face substantial financial burden for COVID-19 hospitalizations, particularly the privately insured.

A strength of this study its use of national, fully-adjudicated claims data. Such data are typically considered complete after a six-month time lag, meaning claims through the latter part of 2020 only became available shortly before the time of writing in May 2021. An additional strength is the inclusion of both privately insured and Medicare Advantage plans. These plans are important sources of coverage for older adults and the elderly,^13,14^ two age groups accounting for high shares of COVID-19 hospitalizations.^15^

This study also has limitations. First, despite strong indirect evidence, we cannot prove that COVID-19 hospitalizations in this study were mostly covered by plans with cost-sharing waivers. Second, if patients did not pay the amounts they were billed, the incidence of actual out-of-pocket spending would differ from the incidence estimated by this study. However, the amount billed to patients still illustrates the financial burden patients may face without cost-sharing waivers. Third, COVID-19 hospitalizations without the diagnosis code for confirmed COVID-19 (U017) were not included. However, hospitals rapidly started using this code during the first half of 2020.^16^ Finally, our database is not necessarily representative of all private and Medicare Advantage plans. However, most privately insured hospitalizations in our study were covered by preferred private organization plans, while most Medicare Advantage hospitalizations were covered by health maintenance organizations, consistent with the national distribution of plan type among privately insured and Medicare Advantage enrollees.^13,17^

## CONCLUSION

Findings suggest that insurer cost-sharing waivers for COVID-19 hospitalizations may not always capture all hospitalization-related care. Moreover, patient financial burden for COVID-19 hospitalizations could be substantial without insurer waivers. The growing trend towards abandonment of these waivers suggests that relying on voluntary insurer actions is not an ideal policy strategy to protect patients from the costs of COVID-19 hospitalizations.^9,10^ To achieve this goal, federal policymakers might consider legislation mandating insurers to waive cost-sharing for COVID-19 hospitalizations throughout the public health emergency.^11^ Such a mandate would ideally include all hospitalization-related care, similar to existing federal mandates which require insurers to fully cover all direct and related costs of COVID-19 tests and vaccines.^12^ Future research should continue to examine patient financial burden of COVID-19 hospitalizations as coverage policies change.

## Data Availability

IQVIA data are proprietary and cannot be shared

## Author Contributions

Dr. Chua had full access to all of the data in the study and takes responsibility for the integrity of the data and the accuracy of the data analysis.

*Study concept and design:* Chua, Conti, Becker

*Acquisition of data:* Chua

*Analysis and interpretation of data:* Chua, Conti, Becker

*Drafting of the manuscript:* Chua

*Critical revision of the manuscript:* Chua, Conti, Becker

*Statistical analysis:* Chua

*Study supervision:* Becker

