## Appendix for "Out-of-Pocket Spending for COVID-19 Hospitalizations in 2020"

**Appendix 1.** Rationale for including hospitalizations with admission and discharge date before instead of through September 30, 2020

Our claims database was considered to be complete through September 30, 2020 at the time we received the data at the end of March 2021. In our database, confinement episodes are defined as a contiguous string of facility room and board claims from the same billing identifier. The first and last dates of this string are the admission and discharge dates, respectively. If this string ends on September 29, 2020, we could be confident there was not another facility room and board claim on September 30, 2020, owing to the completeness of the data. However, if a string includes September 30, 2020, we cannot be sure that there were no other facility room and board claims on October 1, 2020. Thus, we required that the discharge date (last facility room and board claim) be on or before September 29, 2020.

**Appendix 2.** Codes used in analyses

Facility claims were institutional claims with place of service code 21 (hospital) or 23 (emergency department) in which the “billing specialty” variable corresponded to a hospital. These included inpatient hospital services such as room and board, pharmacy, IV therapy, supplies, laboratory, radiology, blood bank, respiratory services, physical/occupational/speech therapy, and inpatient dialysis. These also included facility charges for emergency departments (e.g., revenue codes 0450-0459).

Professional/ancillary claims were defined as one of the following 3 mutually exclusive types of claims:

1) Ambulance claims - place of service code 41-42 (ambulance) OR HCPCS codes between A0000-A9999

2) Clinician claims: (place of service 21 or 23) AND billing specialty variable = clinician

3) Miscellaneous claims: (place of service 21 or 23) AND billing specialty variable = other (including durable medical equipment providers and other facilities, such as dialysis centers)

The four subtypes of clinician claims were defined as follows:

1) Emergency department claims - place of service code 23 AND billing specialty variable = clinician

2) Inpatient evaluation and management claims – place of service code 21 (hospital) AND billing specialty variable = clinician AND procedure codes for clinician services such as initial and subsequent hospital care or hospital discharge services:

99217, 99218, 99219, 99220, 99221, 99222, 99223, 99224, 99225, 99226, 99231, 99232, 99233, 99234, 99235, 99236, 99238, 99239,	99251, 99252, 99253, 99254, 99255, 99291, 99292, 99356, 99357, 99358, 99359, 99360, 99366, 99367, 99368, 99460, 99461, 99462,	99463, 99464, 99465, 99466, 99467, 99468, 99469, 99471, 99472, 99475, 99476, 99477, 99478, 99479, 99480, 99485, 99486, 99487, 99489, 99490, 99491, 99497, 99498, 99499, G0378, G0379, G0390

3) Inpatient diagnostic claims: place of service code 21 (hospital) AND billing specialty variable = clinician AND CPT/HCPCS codes that were a) between 70000-79999 (radiology); b) between 80000-89999 or U0001-U0004 (laboratory tests); or c) CPT codes for the following types of tests:

| **Electrocardiogram** | |
| --- | --- |
| 93000 | Electrocardiogram, routine ECG with at least 12 leads; with interpretation and report |
| 93005 | Electrocardiogram, routine ECG with at least 12 leads; tracing only, without interpretation and report |
| 93010 | Electrocardiogram, routine ECG with at least 12 leads; interpretation and report only |
| 93040 | Rhythm ECG, 1-3 leads; with interpretation and report |
| 93041 | Rhythm ECG, 1-3 leads; tracing only without interpretation and report |
| 93042 | Rhythm ECG, 1-3 leads; interpretation and report only |
| **Vascular diagnostic studies** | |
| 93880 | Duplex scan of extracranial arteries; complete bilateral study |
| 93882 | Duplex scan of extracranial arteries; unilateral or limited study |
| 93886 | Transcranial Doppler study of the intracranial arteries; complete study |
| 93888 | Transcranial Doppler study of the intracranial arteries; limited study |
| 93890 | Transcranial Doppler study of the intracranial arteries; vasoreactivity study |
| 93892 | Transcranial Doppler study of the intracranial arteries; emboli detection without intravenous microbubble injection |
| 93893 | Transcranial Doppler study of the intracranial arteries; emboli detection with intravenous microbubble injection |
| 93895 | Quantitative carotid intima media thickness and carotid atheroma evaluation, bilateral |
| 93922 | Limited bilateral noninvasive physiologic studies of upper or lower extremity arteries, (eg, for lower extremity: ankle/brachial indices at distal posterior tibial and anterior tibial/dorsalis pedis arteries plus bidirectional, Doppler waveform recording and analysis at 1-2 levels, or ankle/brachial indices at distal posterior tibial and anterior tibial/dorsalis pedis arteries plus volume plethysmography at 1-2 levels, or ankle/brachial indices at distal posterior tibial and anterior tibial/dorsalis pedis arteries with, transcutaneous oxygen tension measurement at 1-2 levels) |
| 93923 | Complete bilateral noninvasive physiologic studies of upper or lower extremity arteries, 3 or more levels (eg, for lower extremity: ankle/brachial indices at distal posterior tibial and anterior tibial/dorsalis pedis arteries plus segmental blood pressure measurements with bidirectional Doppler waveform recording and analysis, at 3 or more levels, or ankle/brachial indices at distal posterior tibial and anterior tibial/dorsalis pedis arteries plus segmental volume plethysmography at 3 or more levels, or ankle/brachial indices at distal posterior tibial and anterior tibial/dorsalis pedis arteries plus segmental transcutaneous oxygen tension measurements at 3 or more levels), or single level study with provocative functional maneuvers (eg, measurements with postural provocative tests, or measurements with reactive hyperemia) |
| 93924 | Noninvasive physiologic studies of lower extremity arteries, at rest and following treadmill stress testing, (ie, bidirectional Doppler waveform or volume plethysmography recording and analysis at rest with ankle/brachial indices immediately after and at timed intervals following performance of a standardized protocol on a motorized treadmill plus recording of time of onset of claudication or other symptoms, maximal walking time, and time to recovery) complete bilateral study |
| 93925 | Duplex scan of lower extremity arteries or arterial bypass grafts; complete bilateral study |
| 93926 | Duplex scan of lower extremity arteries or arterial bypass grafts; unilateral or limited study |
| 93930 | Duplex scan of upper extremity arteries or arterial bypass grafts; complete bilateral study |
| 93931 | Duplex scan of upper extremity arteries or arterial bypass grafts; unilateral or limited study |
| 93970 | Duplex scan of extremity veins including responses to compression and other maneuvers; complete bilateral study |
| 93971 | Duplex scan of extremity veins including responses to compression and other maneuvers; unilateral or limited study |
| 93975 | Duplex scan of arterial inflow and venous outflow of abdominal, pelvic, scrotal contents and/or retroperitoneal organs; complete study |
| 93976 | Duplex scan of arterial inflow and venous outflow of abdominal, pelvic, scrotal contents and/or retroperitoneal organs; limited study |
| 93978 | Duplex scan of aorta, inferior vena cava, iliac vasculature, or bypass grafts; complete study |
| 93979 | Duplex scan of aorta, inferior vena cava, iliac vasculature, or bypass grafts; unilateral or limited study |
| 93980 | Duplex scan of arterial inflow and venous outflow of penile vessels; complete study |
| 93981 | Duplex scan of arterial inflow and venous outflow of penile vessels; follow-up or limited study |
| 93985 | Duplex scan of arterial inflow and venous outflow for preoperative vessel assessment prior to creation of hemodialysis access; complete bilateral study |
| 93986 | Duplex scan of arterial inflow and venous outflow for preoperative vessel assessment prior to creation of hemodialysis access; complete unilateral study |
| 93990 | Duplex scan of hemodialysis access (including arterial inflow, body of access and venous outflow) |
| **Echocardiography** | |
| 93303 | Transthoracic echocardiography for congenital cardiac anomalies; complete |
| 93304 | Transthoracic echocardiography for congenital cardiac anomalies; follow-up or limited study |
| 93306 | Echocardiography, transthoracic, real-time with image documentation (2D), includes M-mode recording, when performed, complete, with spectral Doppler echocardiography, and with color flow Doppler echocardiography |
| 93307 | Echocardiography, transthoracic, real-time with image documentation (2D), includes M-mode recording, when performed, complete, without spectral or color Doppler echocardiography |
| 93308 | Echocardiography, transthoracic, real-time with image documentation (2D), includes M-mode recording, when performed, follow-up or limited study |
| 93312 | Echocardiography, transesophageal, real-time with image documentation (2D) (with or without M-mode recording); including probe placement, image acquisition, interpretation and report |
| 93313 | Echocardiography, transesophageal, real-time with image documentation (2D) (with or without M-mode recording); placement of transesophageal probe only |
| 93314 | Echocardiography, transesophageal, real-time with image documentation (2D) (with or without M-mode recording); image acquisition, interpretation and report only |
| 93315 | Transesophageal echocardiography for congenital cardiac anomalies; including probe placement, image acquisition, interpretation and report |
| 93316 | Transesophageal echocardiography for congenital cardiac anomalies; placement of transesophageal probe only |
| 93317 | Transesophageal echocardiography for congenital cardiac anomalies; image acquisition, interpretation and report only |
| 93318 | Echocardiography, transesophageal (TEE) for monitoring purposes, including probe placement, real time 2-dimensional image acquisition and interpretation leading to ongoing (continuous) assessment of (dynamically changing) cardiac pumping function and to therapeutic measures on an immediate time basis |
| 93320 | Doppler echocardiography, pulsed wave and/or continuous wave with spectral display (List separately in addition to codes for echocardiographic imaging); complete |
| 93321 | Doppler echocardiography, pulsed wave and/or continuous wave with spectral display (List separately in addition to codes for echocardiographic imaging); follow-up or limited study (List separately in addition to codes for echocardiographic imaging) |
| 93325 | Doppler echocardiography color flow velocity mapping (List separately in addition to codes for echocardiography) |
| 93350 | Echocardiography, transthoracic, real-time with image documentation (2D), includes M-mode recording, when performed, during rest and cardiovascular stress test using treadmill, bicycle exercise and/or pharmacologically induced stress, with interpretation and report; |
| 93351 | Echocardiography, transthoracic, real-time with image documentation (2D), includes M-mode recording, when performed, during rest and cardiovascular stress test using treadmill, bicycle exercise and/or pharmacologically induced stress, with interpretation and report; including performance of continuous electrocardiographic monitoring, with supervision by a physician or other qualified health care professional |
| 93352 | Use of echocardiographic contrast agent during stress echocardiography (List separately in addition to code for primary procedure) |
| 93355 | Echocardiography, transesophageal (TEE) for guidance of a transcatheter intracardiac or great vessel(s) structural intervention(s) (eg,TAVR, transcatheter pulmonary valve replacement, mitral valve repair, paravalvular regurgitation repair, left atrial appendage occlusion/closure, ventricular septal defect closure) (peri-and intra-procedural), real-time image acquisition and documentation, guidance with quantitative measurements, probe manipulation, interpretation, and report, including diagnostic transesophageal echocardiography and, when performed, administration of ultrasound contrast, Doppler, color flow, and 3D |
| **Electroencephalography** | |
| 95700 | Electroencephalogram (EEG) continuous recording, with video when performed, setup, patient education, and takedown when performed, administered in person by EEG technologist, minimum of 8 channels |
| 95705 | Electroencephalogram (EEG), without video, review of data, technical description by EEG technologist, 2-12 hours; unmonitored |
| 95706 | Electroencephalogram (EEG), without video, review of data, technical description by EEG technologist, 2-12 hours; with intermittent monitoring and maintenance |
| 95707 | Electroencephalogram (EEG), without video, review of data, technical description by EEG technologist, 2-12 hours; with continuous, real-time monitoring and maintenance |
| 95708 | Electroencephalogram (EEG), without video, review of data, technical description by EEG technologist, each increment of 12-26 hours; unmonitored |
| 95709 | Electroencephalogram (EEG), without video, review of data, technical description by EEG technologist, each increment of 12-26 hours; with intermittent monitoring and maintenance |
| 95710 | Electroencephalogram (EEG), without video, review of data, technical description by EEG technologist, each increment of 12-26 hours; with continuous, real-time monitoring and maintenance |
| 95711 | Electroencephalogram with video (VEEG), review of data, technical description by EEG technologist, 2-12 hours; unmonitored |
| 95712 | Electroencephalogram with video (VEEG), review of data, technical description by EEG technologist, 2-12 hours; with intermittent monitoring and maintenance |
| 95713 | Electroencephalogram with video (VEEG), review of data, technical description by EEG technologist, 2-12 hours; with continuous, real-time monitoring and maintenance |
| 95714 | Electroencephalogram with video (VEEG), review of data, technical description by EEG technologist, each increment of 12-26 hours; unmonitored |
| 95715 | Electroencephalogram with video (VEEG), review of data, technical description by EEG technologist, each increment of 12-26 hours; with intermittent monitoring and maintenance |
| 95716 | Electroencephalogram with video (VEEG), review of data, technical description by EEG technologist, each increment of 12-26 hours; with continuous, real-time monitoring and maintenance |
| 95717 | Electroencephalogram (EEG), continuous recording, physician or other qualified health care professional review of recorded events, analysis of spike and seizure detection, interpretation and report, 2-12 hours of EEG recording; without video |
| 95718 | Electroencephalogram (EEG), continuous recording, physician or other qualified health care professional review of recorded events, analysis of spike and seizure detection, interpretation and report, 2-12 hours of EEG recording; with video (VEEG) |
| 95719 | Electroencephalogram (EEG), continuous recording, physician or other qualified health care professional review of recorded events, analysis of spike and seizure detection, each increment of greater than 12 hours, up to 26 hours of EEG recording, interpretation and report after each 24-hour period; without video |
| 95720 | Electroencephalogram (EEG), continuous recording, physician or other qualified health care professional review of recorded events, analysis of spike and seizure detection, each increment of greater than 12 hours, up to 26 hours of EEG recording, interpretation and report after each 24-hour period; with video (VEEG) |
| 95721 | Electroencephalogram (EEG), continuous recording, physician or other qualified health care professional review of recorded events, analysis of spike and seizure detection, interpretation, and summary report, complete study; greater than 36 hours, up to 60 hours of EEG recording, without video |
| 95722 | Electroencephalogram (EEG), continuous recording, physician or other qualified health care professional review of recorded events, analysis of spike and seizure detection, interpretation, and summary report, complete study; greater than 36 hours, up to 60 hours of EEG recording, with video (VEEG) |
| 95723 | Electroencephalogram (EEG), continuous recording, physician or other qualified health care professional review of recorded events, analysis of spike and seizure detection, interpretation, and summary report, complete study; greater than 60 hours, up to 84 hours of EEG recording, without video |
| 95724 | Electroencephalogram (EEG), continuous recording, physician or other qualified health care professional review of recorded events, analysis of spike and seizure detection, interpretation, and summary report, complete study; greater than 60 hours, up to 84 hours of EEG recording, with video (VEEG) |
| 95725 | Electroencephalogram (EEG), continuous recording, physician or other qualified health care professional review of recorded events, analysis of spike and seizure detection, interpretation, and summary report, complete study; greater than 84 hours of EEG recording, without video |
| 95726 | Electroencephalogram (EEG), continuous recording, physician or other qualified health care professional review of recorded events, analysis of spike and seizure detection, interpretation, and summary report, complete study; greater than 84 hours of EEG recording, with video (VEEG) |

4) Other inpatient services, defined as claims with place of service code 21 and billing specialty type = clinician AND procedure codes that were not related to inpatient evaluation and management or diagnostic testing (e.g., anesthesia, surgery, dialysis, and intubation).

**Intensive care utilization**

Intensive care utilization was defined based on revenue codes 0200-0209 (intensive care unit) and revenue codes 0210-0219 (coronary care unit)

**Appendix 3.** Incidence of cost-sharing for facility services among hospitalizations for patients in the lowest quartile of out-of-pocket spending prior to hospitalization

This analysis subset to 3,617 COVID-19 hospitalizations with continuous enrollment between January 2020 and the hospitalization date (1,203 privately insured, 2,414 Medicare Advantage). We summed out-of-pocket spending across medical and pharmacy claims during this period. For each payer type, we calculated the 25^th^ percentiles of this out-of-pocket spending before hospitalization (private insurance: $113.11; Medicare Advantage: $127.37). Among hospitalizations for privately insured patients and Medicare Advantage patients in the lowest quartile, 8.3% and 1.8% had cost-sharing for facility claims. Findings suggests that cost-sharing waivers, rather than patients meeting out-of-pocket maximums, are the primary driver of the low incidence of cost-sharing for facility claims among COVID-19 hospitalizations.
